# Trends in Diabetes Biomarkers and Treatment Statuses of Non-Institutionalized Canadians: Canadian Health Measures Survey 2007 to 2015

**DOI:** 10.1101/2022.05.04.22274698

**Authors:** Yi-Sheng Chao

**Affiliations:** Independent researcher

**Keywords:** Diabetes, pre-diabetes, Canadian Health Measures Survey, biomarker, insulin

## Abstract

**Background:** Diabetes has been a major source of disease burden in Canada. Moreover, untreated diabetes can lead to complications and severe conditions. A few studies exist on the prevalence of diabetes and the adequacy of diabetes management for the Canadian population, and so this study aims to estimate the diabetes prevalence rates using biomarkers and the treatment statuses of non-institutionalized Canadian patients.

**Methods:** The Canadian Health Measures Survey (CHMS) cycles 1 to 4 were conducted between 2007 and 2015 as interviews with non-institutionalized Canadians. Four blood diabetic markers were measured: insulin, glycosylated hemoglobin percentages, random-spot glucose, and fasting glucose. Subjects with levels higher than normal ranges were considered to have pre-diabetes or diabetes. Treatment statuses were categorized into *treated* (using anti-diabetic agents or diagnosed with diabetes), *probably treated* (taking prescriptions or diagnosed with chronic conditions), *potentially treated* (taking any medications or diagnosed with chronic conditions), and *untreated* (not taking any medications and not diagnosed with chronic conditions). Weights were applied to generate nationally representative statistics.

**Results:** The blood insulin levels in cycle 4 were significantly higher than those in cycle 1 (ratio = 1.42, 95% CI = 1.04 to 1.79). The proportions of patients with pre-diabetes and diabetes were estimated differently at 0.75% using random-spot glucose and 42.17% using glycosylated hemoglobin percentages, respectively. The proportions of Canadians with uncontrolled pre-diabetes or diabetes varied from 0.59% using random-spot glucose levels to 4.63% using fasting glucose levels, respectively. Through cycles 1 to 4, the proportions of untreated Canadians with pre-diabetes or diabetes ranged from 3.86% to 3.73%. More than 93% of those with high fasting glucose levels were taking prescription medications or had been diagnosed with chronic conditions (probably treated). Less than 33% of those with high fasting glucose levels were diagnosed or actively being treated with anti-diabetic agents (treated).

**Conclusion:** Diabetes biomarkers might be useful for screening untreated and undertreated patients with pre-diabetes or diabetes. The treatment categories we used indicated different intensities of intervention that might be useful for determining levels of patient outreach and for planning targeted screening in Canada.

## Introduction

In 2017 in Canada, diabetes was the seventh leading cause of death and the third cause of disability.[1] Diabetes Canada predicted more than 5 million or 12.1% of the Canadian population will be diabetic by 2025.[2] In the United States in 2011 and 2012, diabetes was estimated to be prevalent in 12.5% of the American population, and 30% of those with diabetes had not been treated.[3] In Canada, between 2007 and 2011, estimates predicted that 7.55% of Canadian adults were diabetic based on levels of fasting glucose and hemoglobin A1c, and 43.67% of these adults had not been diagnosed.[4] Diabetes is a chronic condition caused by several mechanisms. Type 1 diabetes involves destroyed pancreatic beta cells and insulin deficiency.[5] Type 2 diabetes development is related to genetic predisposition and molecular mechanisms that subsequently lead to impaired beta cells’ glucose responsiveness, insulin resistance, and increases in blood glucose levels.[6, 7] Type 1 diabetes is less frequent[7] and usually occurs at a young age and requires insulin for treatment.[8, 9] Type 2 diabetes develops gradually and is associated with several risk factors, such as being overweight, high blood pressure, and a family history of diabetes.[7] If left untreated and uncontrolled, the long-term complications can include vision loss, cardiovascular disease, and diabetic neuropathy with nerve damage.[7]

Within communities, the detection and treatment of patients with diabetes is challenging. Different diabetes biomarkers, such as fasting and random-spot glucose levels, can yield various estimates of diabetes prevalence. For example, the levels of hemoglobin A1c seem to detect more patients with diabetes, compared to the levels of fasting plasma glucose.[4] Often, after estimating the prevalence of diabetes, some studies do not report on the treatment status of diabetic patients.[4] In other studies, a significant treatment gap exists regarding cardiovascular disease management for diabetic patients.[10] Among patients with hypertension, 14.4% of it was uncontrolled.[11] It remains unclear whether higher proportions of patients with diabetes are treated. Whereas a timely diagnosis and effective treatment are keys to reducing diabetic complications at the individual level,[7] at a population level, it is important to know the numbers of patients with diabetes who are undiagnosed or undertreated. However, continued efforts to identify proportions of undiagnosed or untreated patients with diabetes have been lacking.[4] Since recent trends in diabetes biomarkers have not been reported in Canada, our study fills this gap by providing the trends of diabetes biomarkers and the treatment status of Canadian patients with abnormal diabetes biomarker profiles.

## Methods

Our study uses the data from the biannual Canadian Health Measures Survey (CHMS), cycles 1 to 4 implemented between 2007 and 2015.[12, 13] More than 5,000 non-institutionalized Canadians were sampled for each cycle to derive nationally representative statistics.[12, 13] Institutionalized Canadians and those living on reserves, who constituted less than 4% of Canadians, were excluded from the sampling.[13] Detailed inclusion and exclusion criteria and sampling frames were published elsewhere.[13-16] Participants in the CHMS were interviewed regarding their health conditions and individual characteristics.[13, 17] Mobile units collected blood and urine samples to determine levels of disease biomarkers and environmental chemicals.[18] An exclusive list of biomarkers was published in the CHMS Content Summary.[19]

### Abnormality identification for blood diabetic biomarkers

Since the CHMS screened for levels of biomarkers with a single measurement, it could not replicate the procedures for diagnosing diabetes used in clinical settings, particularly the use of repeated tests and higher thresholds according to the Diabetes Canada diagnosis guidelines.[20] The present study used clinical reference ranges to estimate the proportions of Canadians with levels of biomarkers higher or lower than the higher and lower limits of clinical reference ranges, respectively. The normal ranges for fasting glucose levels were between 3.9 and 6.1 mmol/L or 70 and 110 mg/dL.[21] The upper limit of random glucose levels was 11.1 mmol/L or 200 mg/dL.[22] The range of insulin levels was between 14.35 and 143.5 pmol/L or 2 and 20 μU/mL.[22] The normal range for glycosylated hemoglobin percentages was between 4% and 5.6%.[22]

The CMHS cycles 1 to 4 measured blood insulin levels and glycosylated hemoglobin percentages. Levels of random-spot glucose were measured in cycle 1 and 2, and levels of fasting glucose were measured in cycle 3 and 4.

In the CMHS, the diabetes biomarker measurements used had upper and lower detection limits, so the levels of biomarkers that were higher or lower than these limits could not be quantified. The proportions of Canadian participants with levels lower than the detection levels of insulin were 8.08% and 0.07% in cycle 1 and 2, respectively. There were no participants who had levels of other diabetic measures that were higher or lower than the respective upper and lower detection limits.

### Treatment status identification

*Treatment status* was defined according to the use of medication and the chronic condition diagnoses. This status was estimated by four categories: *treated, probably treated, potentially treated*, and *untreated*. To avoid over-identification, *untreated individuals* were defined as those who participants had not reported any chronic conditions or used the medications listed in Appendix 1. Individuals were regarded as *treated* only if they had been diagnosed with diabetes or were taking diabetic prescription drugs listed in Appendix 1. Some patients diagnosed with diabetes might receive a non-medicinal treatment, such as life-style interventions only.[23]

Individuals were *probably treated* if they were diagnosed with chronic conditions defined by the CHMS (any chronic conditions such as cardiovascular disease, high blood pressure, abnormal lipid profile, stroke, kidney disease, liver disease, thyroid disease, diabetes, cancer, eating disorder, and chronic obstructive pulmonary disease listed in Appendix 1) that might affect clinical biomarkers or were taking any prescription drug. Individuals were regarded as *potentially treated* if they were diagnosed with the above-mentioned conditions or were taking any prescription or over-the-counter drug. The difference between *probably* and *potentially treated* statuses was the use of over-the-counter medications.

*Chronic conditions* were identified using the CHMS data dictionaries and were variables beginning with “ccc.”[24] *Medication use* was based on the Anatomical Therapeutic Chemical (ATC) classification system.[25] Medication listed in the CHMS data was labelled as prescription, over-the-counter drugs, and health products. Over-the-counter drugs and health products were regarded as non-prescription. Treatment status identification using disease reporting and medication has been tried in several studies,[11, 26-28] but the present study adopted multiple categories to demonstrate the uncertainties in determining treatment status.

### Data analysis

The present study used all the CHMS cycles 1 to 4 variables to conduct its trend analysis. [14, 15, 29] First, it summarized all the CHMS variables according to their basic characteristics, such as their maximal and minimal values.[14, 15] Then we matched the survey and bootstrap weights to the variables.[14, 15] Next, we processed the variables to identify missing values and values beyond the detection levels. We also documented the basic characteristics, such as weighted mean, and 25th and 75th percentiles.[14, 15] We conducted the data analysis using R (v3.20)[30] and RStudio (v0.98.113)[31]. Due to the descriptive purpose of the present study, we reported 95% CIs, rather than p values. To protect participant confidentiality and prevent any participants from being identified, Statistics Canada has multiple rules on the types of statistics that can be published. For example, minimal numbers of unweighted sample sizes are required for individual categories.

## Results

The CHMS cycles 1 to 4 estimated that there were 29.2 to 32.3 million non-institutionalized Canadians with diabetes, 50% of whom were female.[14, 15] Through cycles 1 to 4, the mean age was around 39 years.[14, 15] Through these four cycles, the mean BMI remained similar, from 26.0 to 26.2 kg/m^2^ .[14, 15]

### Trends in diabetic markers

The blood insulin levels in cycle 4 were significantly higher than those in cycle 1 (ratio = 1.42, 95% CI = 1.04 to 1.79; mean = 67.78 and 89.87 in cycle 1 and 4, Table 1). The glycosylated hemoglobin percentages in cycle 2 to 4 were not significantly different from the values in cycle 1 in Table 2 (ratio 95% CIs including 1 in cycle 2 to 4, weighted means = 5.58%, 5.69%, 5.41%, and 5.42% in cycle 1 to 4, respectively). Levels of random-spot glucose (mmol/l) were measured in cycle 1 and 2, and the levels in cycle 2 were not significantly different from those in cycle 1 in Table 3 (ratio = 1.09, 95% CI = 0.90 to 1.27, weighted means = 4.96 and 5.08 in cycle 1 and 2, respectively). We measured the levels of fasting glucose in cycles 3 and 4, and the levels in cycle 4 were not significantly different from those in cycle 3 (ratio = 1.03, 95% CI = 0.91 to 1.16, weighted mean = 5.17 and 5.20 in cycle 3 and 4, respectively) (Table 4).

**Table 1.**
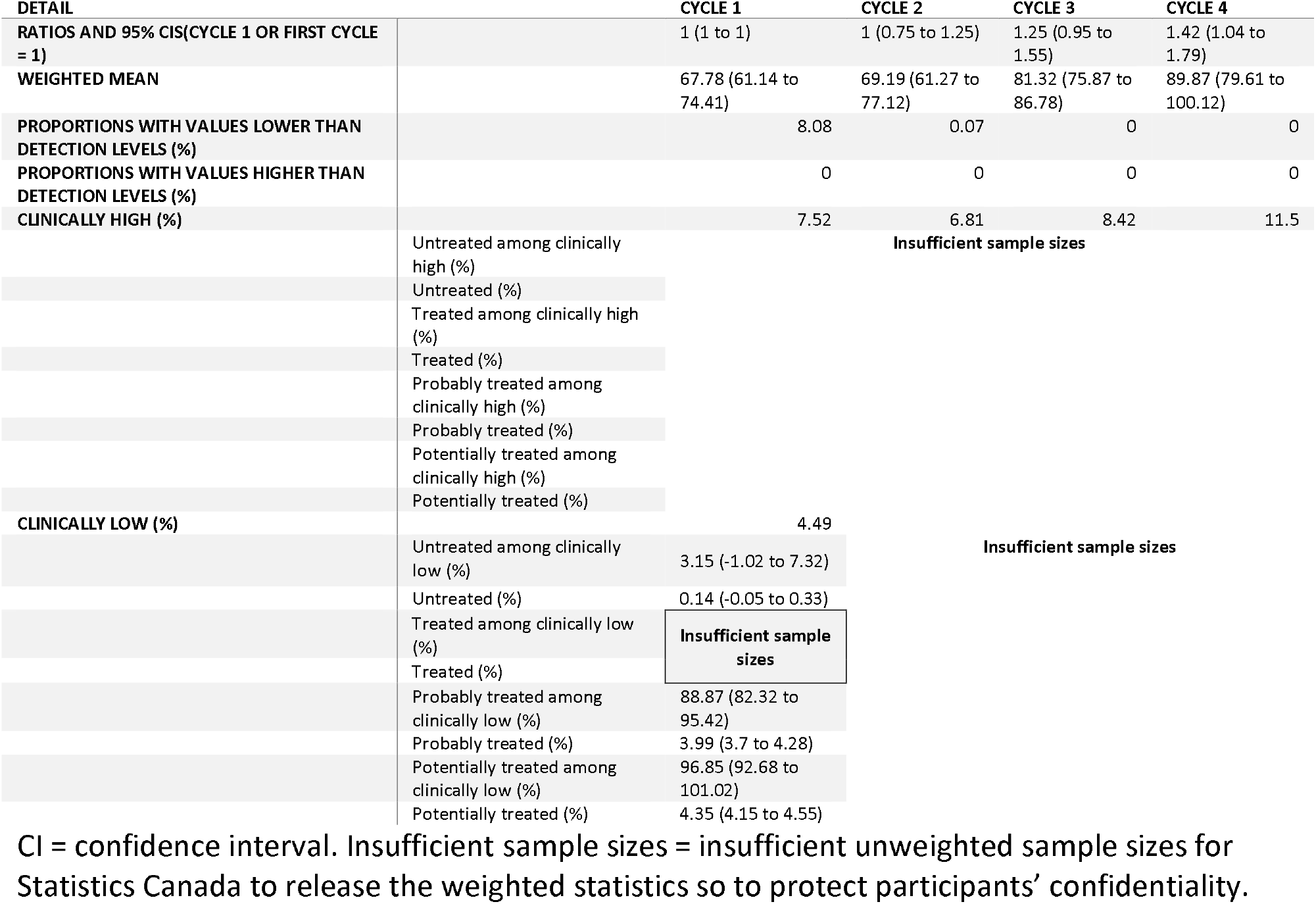
Trends in blood insulin (pmol/l) levels and treatment statuses.

**Table 2.**
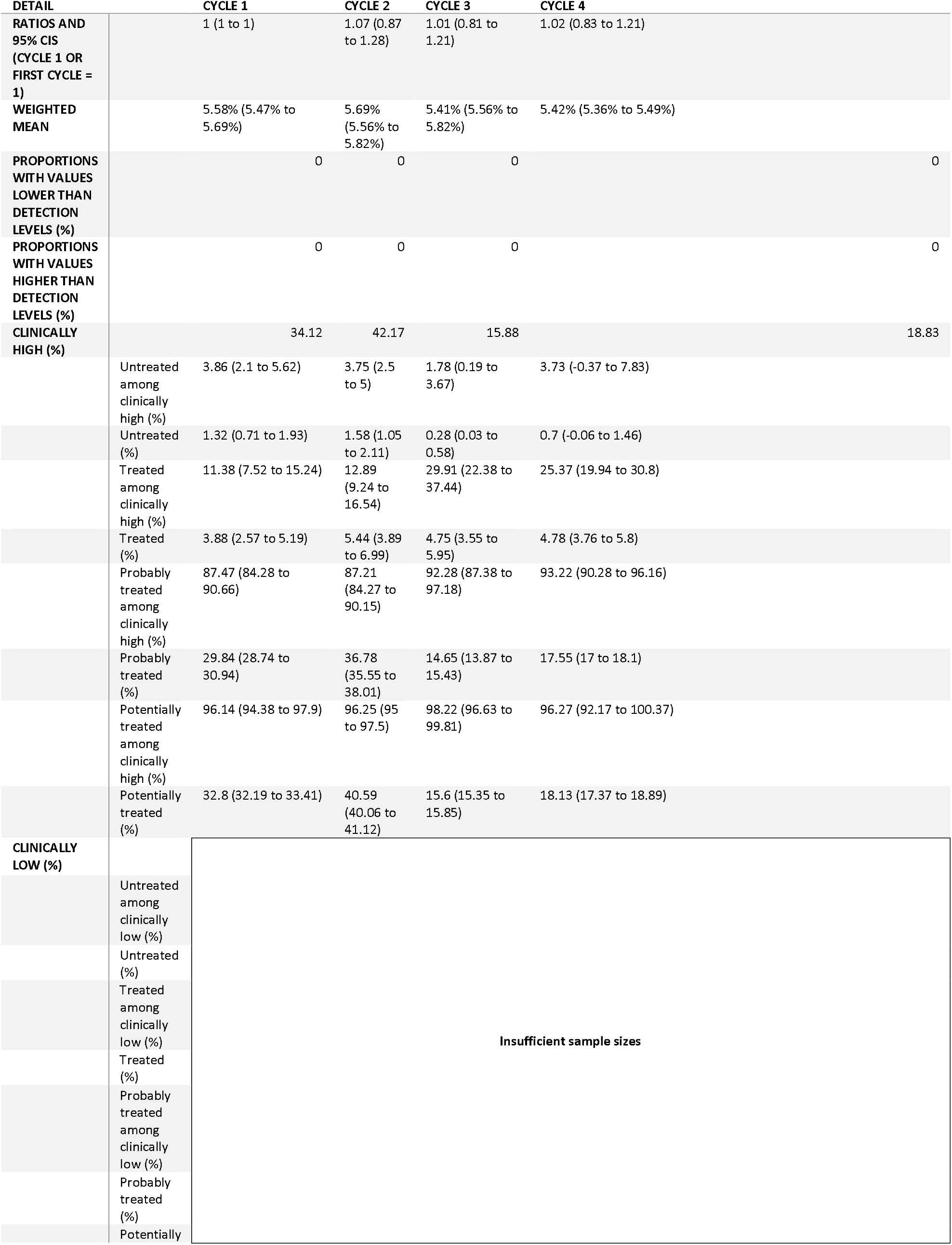
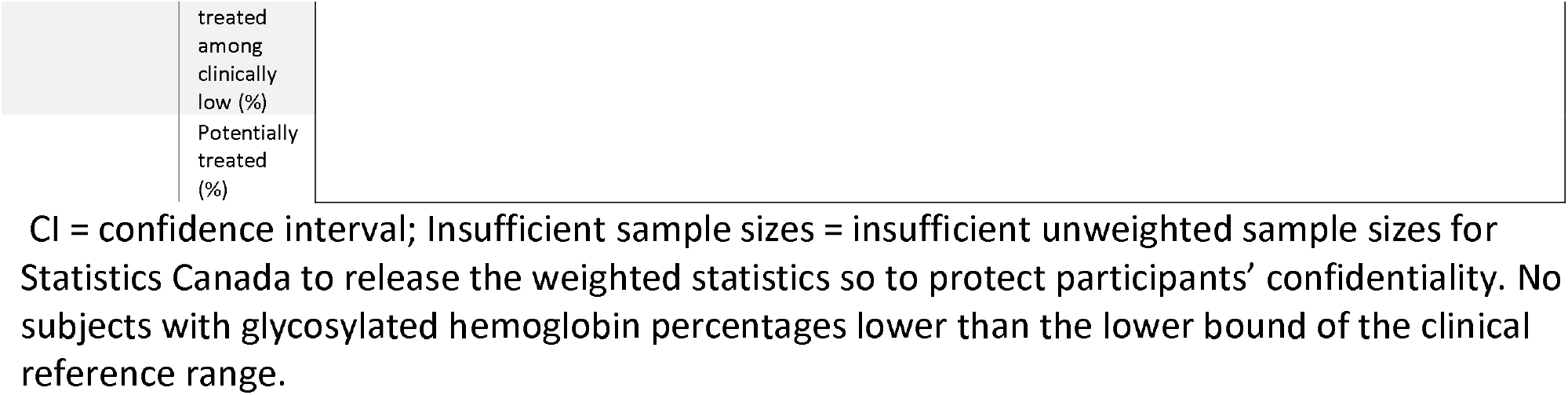
Trends in glycosylated hemoglobin percentages and treatment status.

**Table 3.** Trends in blood random-spot glucose levels (mmol/l) and treatment status.

|  |  | CYCLE 1 | CYCLE 2 | CYCLE 3 | CYCLE 4 |
| --- | --- | --- | --- | --- | --- |
| RATIOS AND 95% CIS (CYCLE 1 OR FIRST CYCLE = 1) |  | 1 (1 to 1) | 1.09 (0.9 to 1.27) | Not<br>measured |  |
| WEIGHTED MEAN |  | 4.96 (4.93 to 5) | 5.08 (4.99 to 5.16) |  |  |
| PROPORTIONS WITH VALUES LOWER THAN<br>DETECTION LEVELS (%) |  | 0 | 0 |  |  |
| PROPORTIONS WITH VALUES HIGHER THAN<br>DETECTION LEVELS (%) |  | 0 | 0 |  |  |
| CLINICALLY HIGH (%) |  | 0.75 | 0.84 |  |  |
|  | Untreated among clinically high (%) | 0 | 0 |  |  |
|  | Untreated (%) | 0 | 0 |  |  |
|  | Treated among clinically high (%) | 78.84 (71.24 to<br>86.44) | 93.02 (84.55 to<br>101.49) |  |  |
|  | Treated (%) | 0.59 (0.53 to<br>0.65) | 0.78 (0.7 to 0.86) |  |  |
|  | Probably treated among clinically<br>high (%) | 100 (100 to 100) | 100 (100 to 100) |  |  |
|  | Probably treated (%) | 0.75 (0.75 to<br>0.75) | 0.84 (0.84 to 0.84) |  |  |
|  | Potentially treated among clinically<br>high (%) | 100 (100 to 100) | 100 (100 to 100) |  |  |
|  | Potentially treated (%) | 0.75 (0.75 to<br>0.75) | 0.84 (0.84 to 0.84) |  |  |
| CLINICALLY LOW (%) |  | Insufficient sample sizes |  |  |  |
|  | Untreated among clinically low (%) |  |  |  |  |
|  | Untreated (%) |  |  |  |  |
|  | Treated among clinically low (%) |  |  |  |  |
|  | Treated (%) |  |  |  |  |
|  | Probably treated among clinically<br>low (%) |  |  |  |  |
|  | Probably treated (%) |  |  |  |  |
|  | Potentially treated among clinically<br>low (%) | Insufficient sample sizes |  |  |  |
|  | Potentially treated (%) |  |  |  |  |

CI = confidence interval; Insufficient sample sizes: insufficient unweighted sample sizes for Statistics Canada to release the weighted statistics so to protect participants' confidentiality.

**Table 4.**
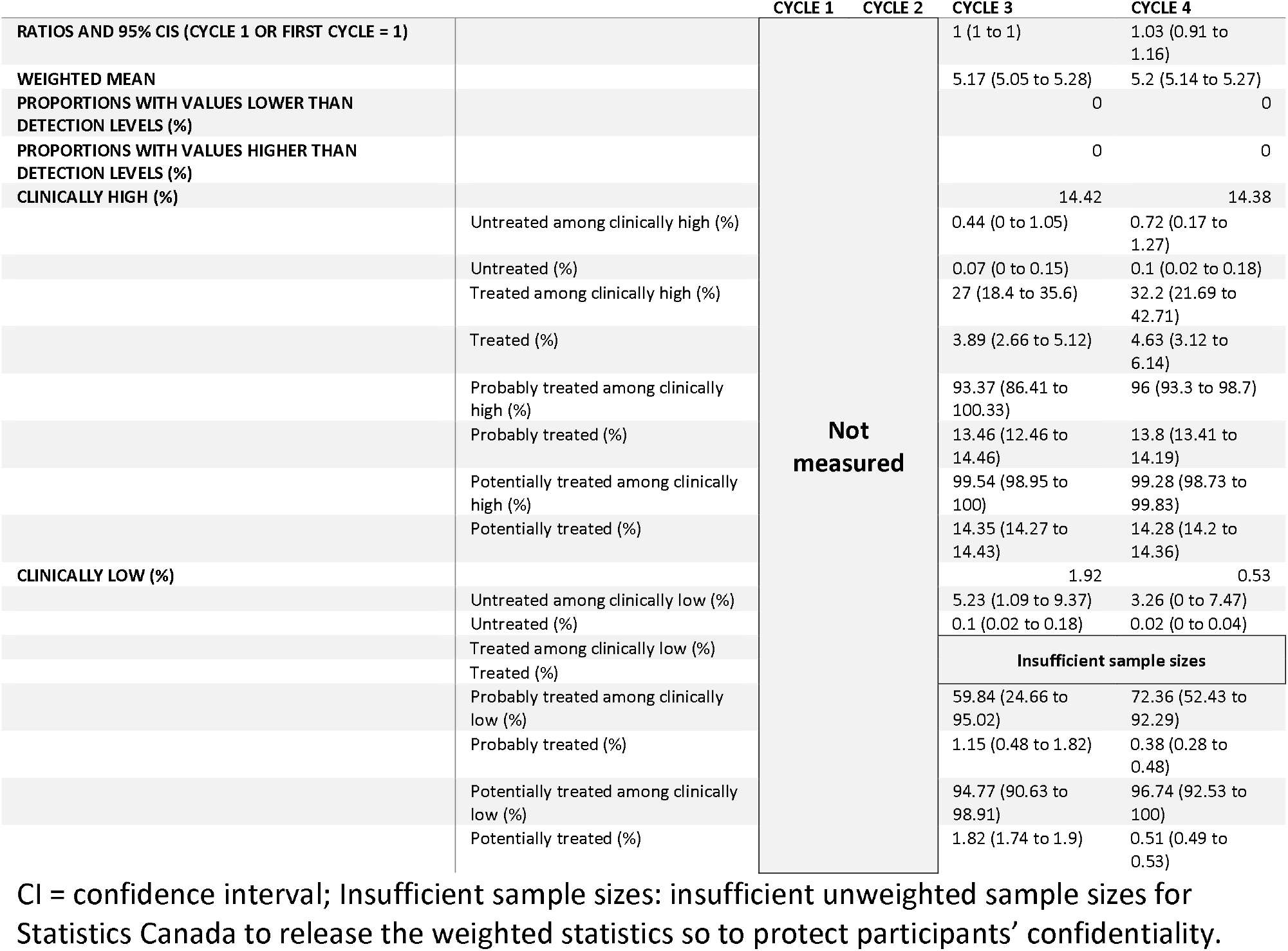
Trends in blood fasting glucose levels (mmol/l) and treatment status.

### Proportions with levels higher or lower than reference levels

The proportions of the population with levels higher or lower than the clinical reference ranges varied across the CHMS cycles and diabetes biomarkers. The proportions with insulin levels higher than the reference ranges were 7.52%, 6.81%, 8.42%, and 11.5% in the CHMS cycles 1 to 4, respectively (Table 1). Only 4.49% of Canadians were found to have insulin levels lower than the clinical reference ranges in cycle 1. The unweighted sample sizes in cycles 2 to 4 were not large enough for Statistics Canada to release weighted statistics. The proportions of the population with high glycosylated hemoglobin were 34.12%, 42.17%, 15.88%, and 18.83% in cycles 1 to 4, respectively (Table 2). The proportions with high random-spot glucose levels were 0.75% and 0.84% in cycles 1 and 2, respectively (Table 3). The proportions with high fasting glucose levels were 14.42% and 14.38% in cycles 3 and 4, respectively (Table 4).

The lower limits of the reference ranges for random-spot glucose levels were not defined and not available (Table 3). No subjects were identified with low glycosylated hemoglobin percentages in cycles 1 to 4 (Table 2). The proportions with low fasting glucose levels were 1.92% and 0.53% in cycles 3 and 4, respectively (Table 4).

### Treatment status

Based on Statistics Canada’s regulations, no sufficient unweighted sample sizes were available to determine the proportions of various treatment statuses for high insulin and low fasting glucose levels. There were no subjects with low glycosylated hemoglobin percentages.

In cycle 1, among those with low insulin levels, 3.15%, 88.87%, and 96.85% were untreated, probably treated, and potentially treated, respectively (Table 1). However, no sufficient unweighted subjects were available to determine the proportions of those treated. Among those with high levels of glycosylated hemoglobin percentages, the proportions of those who were untreated were 3.86%, 3.75%, 1.78%, and 3.73% in cycles 1 to 4, respectively. The proportions of those treated ranged from 11.38% to 29.91% across cycles 1 to 4. The proportions of those probably treated ranged from 87.21% to 93.22% across cycles 1 to 4. Among those with high insulin levels who were potentially treated, the range was 96.14% to 98.22%.

Among those with high random-spot glucose levels, no untreated subjects were identified, and the proportion of those who were probably or potentially treated was 100% across cycles 1 to 2. The proportions of those treated were 78.84% and 93.02% in cycles 1 and 2, respectively. Among those with high fasting glucose levels, the proportions of those who were untreated were 0.46% and 0.72% in cycles 1 and 2, respectively. The proportions of those being treated were 27.0% and 32.2% in cycles 1 and 2, respectively. The proportions of those being probably or potentially treated were more than 93.37% in cycles 1 and 2. Among those with low fasting glucose levels, 5.23% and 3.26% were untreated in cycles 3 and 4, respectively. The proportions of those being probably treated were 59.84% and 72.36% in cycles 3 and 4, respectively. The proportions of those being potentially treated were 94.77% and 96.74% in cycles 3 and 4, respectively.

### Overall proportions of patients with pre-diabetes or diabetes identified

Among all non-institutionalized Canadians, the proportions of untreated patients with diabetes and pre-diabetes identified by the biomarkers varied. Insulin levels failed to identify enough diabetic patients for Statistics Canada to release the estimates. Glycosylated hemoglobin percentages identified 3.86% to 3.73% of untreated Canadians with diabetes or pre-diabetes throughout cycles 1 to 4. Random-spot glucose levels failed to identify any untreated diabetic patients. Fasting glucose levels identified 0.07% and 0.1% of Canadians with diabetes and those who were untreated for diabetes, respectively.

In contrast, the proportions of patients with diabetes being treated also varied. Since these patients were treated and their biomarker levels remained high, they could be considered patients with uncontrolled or poorly controlled diabetes. Glycosylated hemoglobin percentages identified 3.88% to 4.78% of Canadians with uncontrolled pre-diabetes or diabetes through cycles 1 to 4. Random-spot glucose levels identified 0.59% to 0.84% of Canadians with uncontrolled pre-diabetes or diabetes in cycles 1 and 2, respectively. Fasting glucose levels identified 3.89% and 4.63% of Canadians with uncontrolled pre-diabetes or diabetes in cycles 3 and 4, respectively.

## Discussion

Through cycles 1 to 4, blood insulin levels among non-institutionalized Canadians increased significantly. The proportions of Canadians with high or low levels of diabetes biomarkers varied across the CHMS cycles and the types of diabetes biomarkers. The proportions of treatment also varied depending on the biomarkers and treatment indicators. Previous research on the treatment status of patients with diabetes is scarce and is not representative of the Canadian population.[32] Thus, the trend analysis of diabetes markers is feasible and has important implications for the detection of patients with diabetes in populations. First, across the four CHMS cycles conducted between 2007 and 2015, except for blood insulin levels, the levels of diabetes biomarkers, blood glycosylated hemoglobin percentages, random-spot glucose, and fasting glucose did not change significantly, compared to the levels first measured among non-institutionalized Canadians.[14, 15] However, the lack of significant changes over time should be further examined with other statistics. For example, the proportions of non-institutionalized Canadians taking anti-diabetic agents increased from 3.45% to 4.07% from cycles 1 to 4.[15] The proportions of being diagnosed with diabetes also increased from 3.97% to 5.27% from cycles 1 to 4.[15] Whether this finding will be identified in the next CHMS cycles needs to be confirmed.

Second, the proportions of patients with pre-diabetes or diabetes identified by diabetes biomarkers vary widely, from less than 1% by random-spot glucose levels to more than 40% by glycosylated hemoglobin percentages. The upper limits of random-spot glucose are set to a high level at ≥11.1 mmol/L or ≥200 mg/dL that is applicable to patients with symptoms of hyperglycemia.[22] Without information on the symptoms of hyperglycemia, an application of this standard detects small proportions of diabetic patients in the population at 0.75% and 0.84% in cycles 1 and 2, respectively. In contrast, using glycosylated hemoglobin percentages, the proportions of patients with pre-diabetes or diabetes are estimated to be as high as 42.17%. In contrast, based on biomarkers, diabetes prevalence in the general population in the United States was estimated to be 6.5%.[33] It is unclear whether using 5.6% glycosylated hemoglobin to screen for patients with pre-diabetes and diabetes may result in over-identification. Diabetes identification using this biomarker in the general population needs further research.

Third, four categories of treatment statuses can be used to guide the efforts to identify untreated or undertreated patients. Being identified as *probably* or *potentially* treated suggests different levels of exposure to health care systems. The diagnosis of chronic conditions documented in the CHMS has been used to assign a *probably treated* status. A current use of prescription or over-the-counter medications can lead to the identification of a *potentially treated* status. Based on such information, more than 93% of those with high fasting glucose levels were taking prescription medications or had been diagnosed with chronic conditions, whereas less than 33% of them were actively treated with anti-diabetic agents. This suggests that about 60% of those with high fasting glucose levels could be identified and treated medically, if they were not experiencing life-style management for diabetes or undergoing treatment for other conditions. Those with a *potentially treated* status without definite diabetes treatment could have been detected if screening was available where they obtained medications. The proportions of these treatment statuses can guide health care decision makers as to how and where to approach untreated patients with diabetes.

Last, we need to focus on the patients with diabetes, who do not have a clear indication for receiving medical treatment. Using glycosylated hemoglobin percentages or fasting glucose levels, the proportions of untreated Canadians with pre-diabetes or diabetes ranged between 0.7% and 1.58% or 0.07% and 0.1%, respectively. These estimates were lower than those reported in the US[3] or the rates between 2007 and 2011 in Canada.[4] To what extent the differences can be attributed to prevalence or methodological differences remains unclear. However, despite these differences, important economic and health consequences exist for individuals and the public with respect to delaying diabetes treatment.[34] If biomarker screening is considered for untreated diabetes, more precise estimates of the numbers of affected patients and the associated costs and barriers to screening will be major issues. Future releases of CHMS data may help to provide more accurate estimates.

Diabetes screening may be in a greater need during and after the coronavirus disease 2019 (COVID-19) pandemic that has reduced individual contact and the exposure to health care systems.[35] Co-morbidity, especially diabetes, is associated with severe COVID-19 symptoms and more deaths.[36] Early detection of diabetes with biomarkers can help to prevent disease progression and complications.[37] Thus, the detection of diabetes will play a more important role in health care in Canada and elsewhere.

### Limitations

This analysis has several limitations despite the use of national representative data, multiple cross-sectional data, and decreasing proportions of subjects with levels lower than the detection limits over time. The CHMS is a biomonitoring study that aimed to provide descriptive statistics but had no power analysis for any specific hypotheses. To be consistent with a previous study, reference ranges were chosen as such.[4] Other reference ranges adopted by professional societies might lead to some differences in the proportions of patients with diabetes.[20] The CHMS surveyed non-institutionalized Canadians[13-15], and its results were not likely to be applicable to those institutionalized. Moreover, the biomarker levels were not adjusted for the effects of glucose-lowering agents. In addition, the biomarker levels were not adjusted for population structures. Regarding the glycosylated hemoglobin percentages, the diagnosis of pre-diabetes was not separated from diabetes. The measures to prevent the spread of COVID-19 also denied our access to the CHMS, so we were unable to retrieve further research on details, such as the proportions of diabetes based on the glycosylated hemoglobin percentages.

## Conclusion

The levels of three diabetes biomarkers—glycosylated hemoglobin percentages, random-spot glucose, and fasting glucose—for non-institutionalized Canadians remained stable between 2007 and 2015. Insulin levels were significantly higher in cycle 4 than in cycle 1. The proportions of patients with pre-diabetes or diabetes were estimated differently, from 0.75% using random-spot glucose to 42.17% using glycosylated hemoglobin percentages. The use of anti-diabetic agents was used to identify treated patients with diabetes, and the proportions varied from 0.59% using random-spot glucose levels to 4.63% using fasting glucose levels. The use of prescription medications and the diagnoses of the chronic conditions documented in the CHMS were used to identify those who might be probably treated for pre-diabetes or diabetes. The proportions of probable treatment ranged from 0.75% using random-spot glucose levels to 36.78% using glycosylated hemoglobin percentages. Those without chronic conditions or medication use were considered untreated. The proportions of untreated patients with diabetes ranged from 0% using random-spot glucose levels to 1.58% using glycosylated hemoglobin percentages. The use of diabetes biomarkers to screen for patients with pre-diabetes or diabetes might yield unexpected results, and so this screening needs to be planned well. For example, random-spot glucose levels might not be useful for identifying patients with diabetes, and glycosylated hemoglobin percentages might identify high proportions of patients with diabetes in the population. In addition, the treatment categories used in the present study might be useful for planning targeted screening in different contexts.

## Data Availability

It is against the policy of Statistics Act of Canada to release the CHMS data. The collection of CHMS data has been approved by the ethics committee within the governments of Canada. The CHMS data have been de-identified and maintained by Statistics Canada. The data can be accessed through the Research Data Centres administered by Statistics Canada. The details and eligibility for obtaining data access can be found online at https://www.statcan.gc.ca/eng/rdc/process.

## Declaration

## Acknowledgement

The analysis presented in the present study was conducted at the Quebec Interuniversity Centre for Social Statistics (QICSS), which is part of the Canadian Research Data Centre Network (CRDCN). The services and activities provided by the QICSS are made possible by the financial or in-kind support of the Social Sciences and Humanities Research Council (SSHRC), the Canadian Institutes of Health Research (CIHR), the Canada Foundation for Innovation (CFI), Statistics Canada, the Fonds de recherche du Québec and the Quebec universities. The views expressed in this paper are those of the authors, and are not necessarily those of the CRDCN or its partners. The authors thank Dr To-Pang Chen for his constructive comments.

Not applicable.

## Ethics approval

This secondary data analysis was approved by the ethics review committee at the Centre Hospitalier de l’Université de Montréal. All methods were performed in accordance with the guidelines and regulations relevant to the analysis of public data.

## Consent to participate

Written informed consent for the Canadian Health Measures Survey was obtained by Statistics Canada and is not accessible to researchers. Due to this restriction, the requirement of written informed consent was waived by the ethics review committee at the Centre Hospitalier de l’Université de Montréal.

## Author contribution

YSC conceptualized and designed this study, managed and analyzed data, and drafted the manuscript.

## Consent for publication

Participants’ consents for publication are not required for this data analysis project.

